# Late start of antenatal care in Burundi: Effect of socio-demographic, socio-cultural, and health system-related factors

**DOI:** 10.1101/2023.12.12.23299879

**Authors:** Jean M. Butoyi, Caleb K. Sagam, Belyse Munezero, Anatole Nkeshimana, Yves Coppieters

## Abstract

Despite intensive international efforts to extend the coverage of primary healthcare services for women, pregnancy and childbirth still represent a high-risk period for both mother and child, particularly in low- and middle-income countries. Lack of access to antenatal care and its inadequate use during pregnancy contribute to maternal mortality. This study aimed to establish the factors associated with late start of antenatal care (ANC) in Muramvya health district, Burundi. A cross– sectional study was carried out in six randomly selected health facilities of the Muramvya district. The study population consisted of pregnant women who attended antenatal clinics. A total of 280 women were included in the study. Data were collected through face-to-face interviews. Descriptive statistics, bivariate and multivariate logistic regression models were used to measure the determinants of the late start of ANC after 12 weeks of gestation. ANC was initiated after 12 weeks of pregnancy in 72.50% of women, 58.6% and 13.9% respectively in the second and last trimester. Based on Multivariate logistic regression: Women between 20 and 35 years of age have a lower risk of starting ANC late than adolescents (AOR = 0.18; 95%CI:0.03-0.83). Lack of knowledge about the interventions offered during ANC (AOR = 2.37; 95%CI:1.32-4.25), late recognition of pregnancy status (AOR= 6.7; 95%CI:2.41-18.66), unplanned pregnancy (AOR = 1.85 95%CI:1.01-3.40) and the time for women to reach the health facility: between 30minutes to 1 hour (AOR = 2.40 95%CI:1.25-4.59) and more than 1 hour of walking (AOR = 2.15; 95%CI:1.01-4.60) are associated with the late start of ANC. The results of the study show that most women start ANC late. There is a need for effective behavioral interventions and public awareness and education targeting adolescents and women to improve early initiation of ANC

## INTRODUCTION

Reducing maternal and infant mortality remains a priority on the global political agenda for development, as demonstrated by its inclusion in sustainable development goal 3.1(1). Despite intensive international efforts to extend the global coverage of primary healthcare services for women, pregnancy and childbirth still represent a high-risk period for both mother and child, particularly in low- and middle-income countries(2).

Infections undetected during pregnancy, such as malaria, syphilis, tuberculosis, tetanus or human immunodeficiency virus (HIV)/acquired immunodeficiency syndrome (AIDS) infection, as well as high blood pressure, diabetes and others often complicate or aggravate pregnancy and pose a significant risk to both mother and child(2). Nearly 2.3 million newborns die every year in their first month of life, mainly in low- and middle-income countries(3). Around 800 women die every day worldwide from preventable causes associated with pregnancy and childbirth, and 95% of these deaths occur in low- and middle-income countries, over 70% of them in sub-Saharan Africa;. maternal mortality ratio in 2020 was 430 maternal deaths per 100,000 live births, compared with 12 maternal deaths per 100,000 live births in developed countries(4).

To address this situation, one of the pillars of reducing maternal and infant morbidity and mortality is antenatal care (ANC)(5). In 2016, the World Health Organization (WHO) recommended at least eight antenatal care visits. It is generally acknowledged that lack of access to antenatal care and its and poor utilization of ANC during pregnancy contribute to maternal mortality(6,7). Antenatal consultation ensures that risk factors associated with pregnancy are screened, and gives better results if it is carried out early and repeated at regular time intervals(8).

Although recommended by the WHO, early ANC remains a challenge in developing countries, only 52% in low- and middle-income countries, compared with more than 80% in developed countries(8,9). In East Africa, ANC coverage remains well below 50% in most countries: 29% in Uganda (DHS 2016), 29% in Kenya (DHS 2022), 24% in Tanzania (DHS 2016), except Rwanda where it is 59% (DHS 2020) (10).

In Burundi, maternal and neonatal mortality remains high, at 334 maternal deaths per 100,000 live births and 23 neonatal deaths per 1,000 live births respectively(11). These results are still far from the targets of the third sustainable development goal, set at less than 70 maternal deaths for the maternal mortality ratio, with no country in the world exceeding 2 times the global target, and less than 12 deaths for neonatal mortality(1). The objective of this study was to establish the socio-demographic, socio-cultural, and pregnancy-related and health system-related factors that influence start of ANC among pregnant women seen for ANC in Muramvya health district.

## METHODS

### Participants and setting

The study area was Muramvya health district. This is one of two health districts in the Muramvya health province. The district’s health system comprises a district office, a district hospital and 18 health centers, 12 of which are public or faith-based and 6 private. ANC services are offered in all public and faith-based health centers. The study population consisted of pregnant women who attended antenatal clinics in 6 of the 12 public and religious health centers in the Muramvya district. All pregnant women attending ANC who knew the date of their last menstrual period and who agreed to participate freely in the study were included. Women who were from outside the study area and those who did not consent were excluded.

### Design and data collection

This was a cross-sectional quantitative survey done in November 2019 to 20 March 2020. The questionnaire consisted of closed questions and a few open questions. The questions were asked in Kirundi, as this is the language spoken by all Burundians. French was used for those women who understood and wished to use it. Data collection took place at each health center in the study site; before starting the interview, the interviewer introduced himself to the health center manager to announce his presence. Pregnant women were interviewed before being seen for a consultation. The questionnaire lasted an average of 15 minutes. At the end of each week, the questionnaires were collected and checked for completeness and correct completion.

### Data analysis

After data collection, all questionnaires were double-checked to ensure that the information collected was complete, legible and consistent. Epi info software version 7.2.3.0 was used to create the data entry mask and record the data. The data was exported to Stata 15.0 for analysis. The results of the analysis were considered statistically significant for a p value <0.05. Quantitative data were described according to their medians and interquartile ranges. Frequency tables were used to describe categorical data.

To investigate the association between time to onset of ANC and the independent variables, bivariate analysis was performed using Pearson’s chi-square test. The direction, degree of statistical significance and strength of the association between an independent variable and the variable of interest were measured by the OR and 95% confidence interval. This allowed the identification of potential predictors of late onset of ANC.

All variables with a p <=0.20 following the bivariate analysis were included in the multivariate logistic regression model to adjust the variables and control for confounding factors. To obtain a parsimonious model, the backward variable selection method was used. The Akaike information criterion (AIC) was used to select the best model (the one with a low AIC). Potential predictors of late onset of ANC according to pregnant women seen in ANC were identified. Adjusted ORs with their 95% confidence intervals were calculated and interpreted. To determine the validity of the model: its discriminatory power was determined using the ROC curve, the Hosmer-Lemeshow test was performed to check the model’s adequacy, and the Linktest was used for model specification.

### Ethical aspects and protection of privacy

This study was conducted according to the guidelines laid down in the Declaration of Helsinki. Approval was sought from National Institute of Public Health (NIPH)institutional ethics committee to conduct the study. Written consent was sought from the study participants who participate in the study voluntarily and those who didn’t accept to participate were not coerced / discriminated against in any way. Ethical principles on justice, autonomy, beneficence and confidentiality were adhered to. Information collected was kept securely and confidentiality was maintained throughout the study. The data collected did not have any personal identifiers as unique codes generated by the researchers were assigned to each participant. No harm was done to research participants as the research was non-invasive.

## RESULTS

### Description of the sample

Two hundred and eighty (280) pregnant women attending antenatal clinics were surveyed. The median age of the participants was 29 years (range17-47, IQR 24-33). More than half of the women (58.6%) had 2 children. Antenatal consultations were initiated late (>12 weeks of pregnancy) in 72.5% of pregnant women, including 58.6% and 13.9% of women in their second and last trimesters respectively. Antenatal care was done early in 27.5% of the participants. Gestational age at the start of the first antenatal visit ranged from 1 month (4 weeks) to 9 months (40 weeks). The median gestational age was 16 weeks (IQR 12-21).

### Socio-demographic characteristics of participants

Table 1 shows that 75.7% of participants are in the 20-35 age group, 91.8% are married, 56.8% had primary education and 90.4% are farmers with a monthly income of between BIF 20,000 and 40,000 (44.3%). Their husbands have primary education (62.1%) and are generally farmers (75.7%). The households in which these pregnant women live consist of five or more people (48.6%).

**Table 1:**
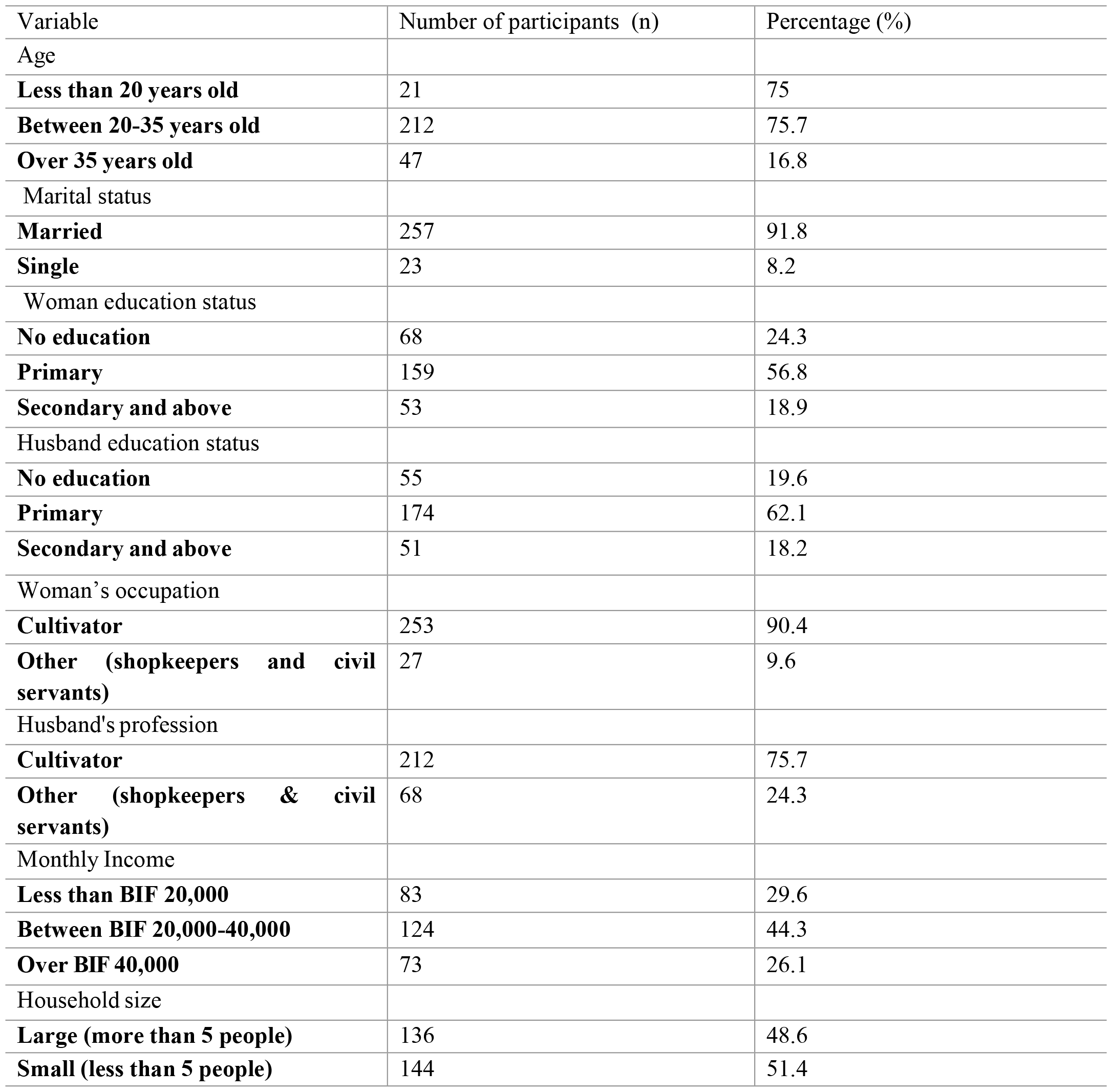
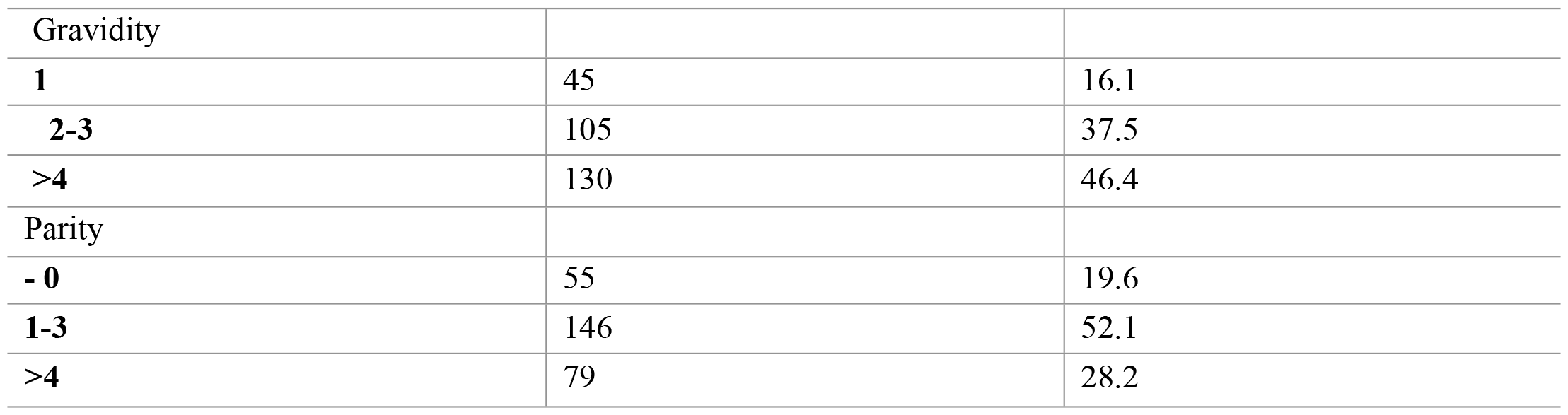
Socio-demographic characteristics of respondents.

### Socio-cultural characteristics of participants

**Table 2:** Socio-cultural characteristics of respondents.

| Variable | Number of participants (n) | Percentage (%) |
| --- | --- | --- |
| Media information (radio) |  |  |
| **Yes** | 166 | 59.3 |
| **No** | 114 | 40.7 |
| Knowledge of ANC |  |  |
| **Little knowledge** | 150 | 53.6 |
| **Better knowledge** | 130 | 46.4 |
| Benefits of doing ANC |  |  |
| **Yes** | 183 | 65.4 |
| **No** | 97 | 34.6 |
| Decision by the woman about ANC |  |  |
| **herself** | 214 | 76.4 |
| **Other (spouse, mother-in-law)** | 66 | 23.6 |
| Encouragement from husband/ANC |  |  |
| **Yes** | 238 | 85.0 |
| **No** | 42 | 15.0 |

The pregnant women without a radio (40.7%). The participants (53.6%) did not mention any or only one intervention offered during ANC and 34.6% did not know that early ANC was beneficial. When deciding to undergo ANC, 76.4% of the women were autonomous and 85.0% were encouraged by their husbands.

### Characteristics related to pregnancy and those related to the care system, according to the participants

**Table 3:**
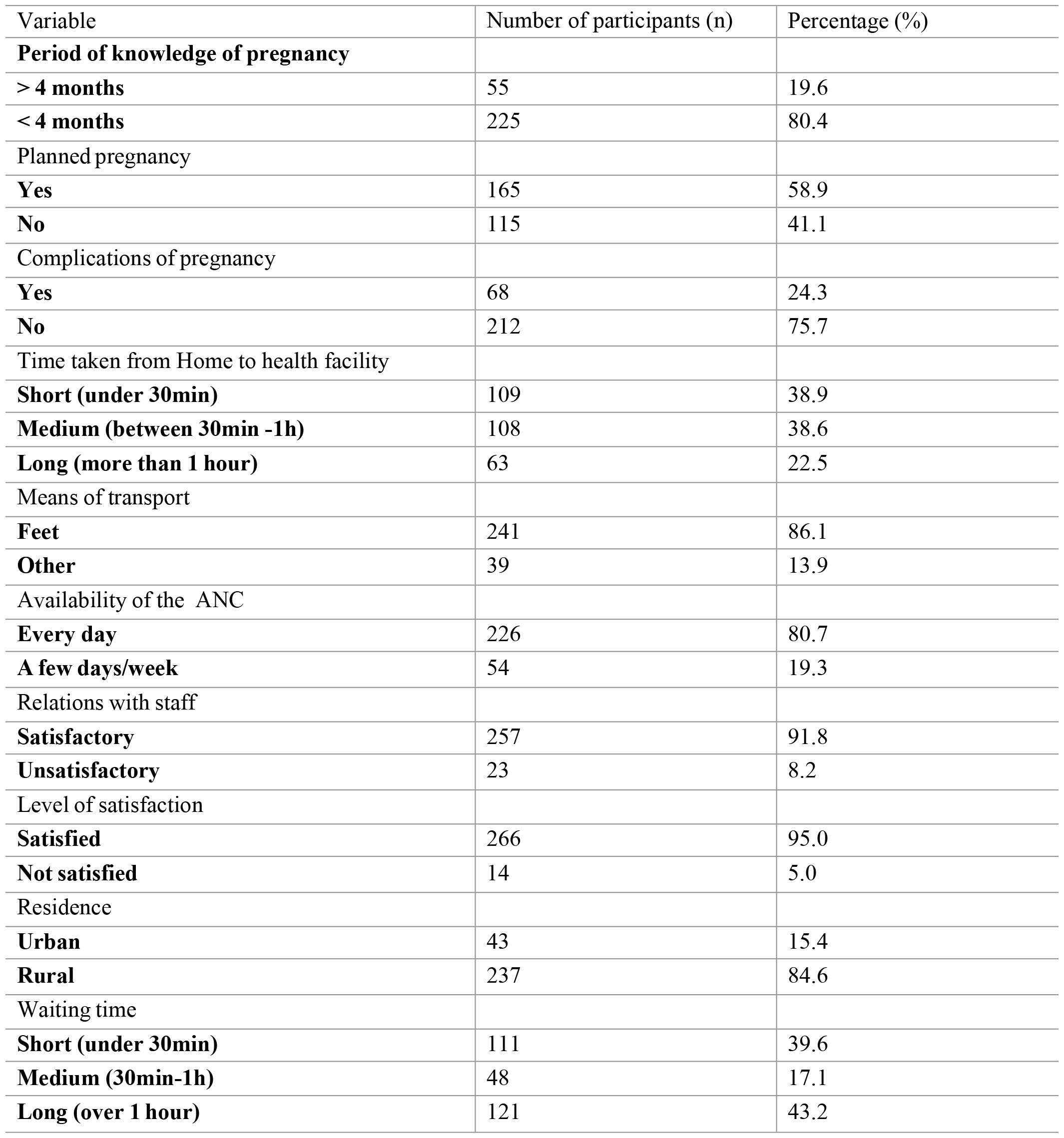
Pregnancy-related and healthcare system-related characteristics of respondents.

Pregnant women (80.4%) knew they were pregnant before 4 months of gestational age, the pregnancy was planned for 58.9% and no complications were observed for 75.7%. 84.6% of the women lived in rural areas. Pregnant women (86.1%) walk to the health facility, and 22.5% of them walk for more than 1 hour.

ANC services are available every day (80.7%) and the relationship they have with health facility staff is considered satisfactory (91.8%). 43.2% of pregnant women waited more than an hour before being seen by health facility staff.

### Relationship between socio-demographic characteristics of pregnant womenand late start of ANC

**Table 4:**
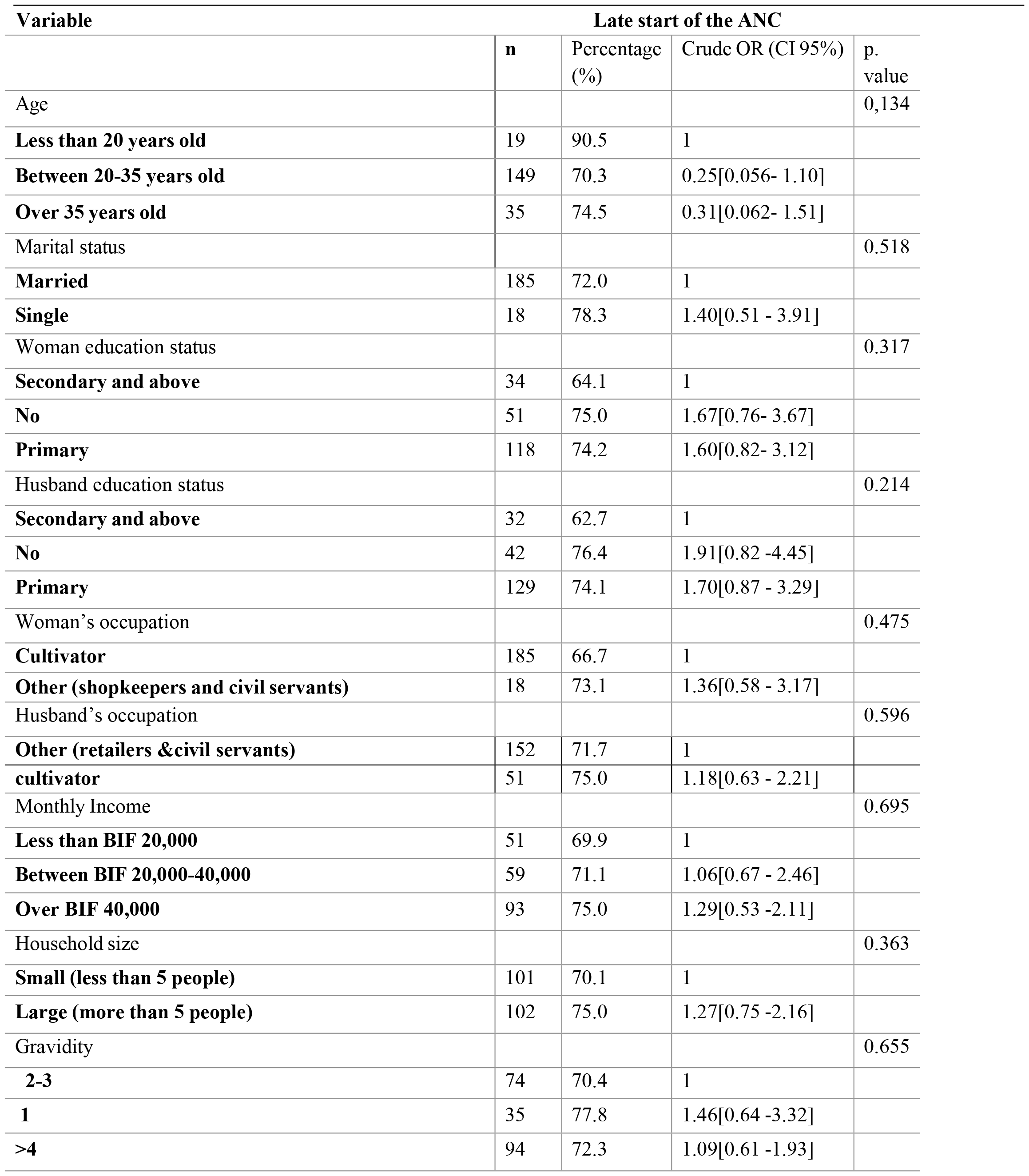
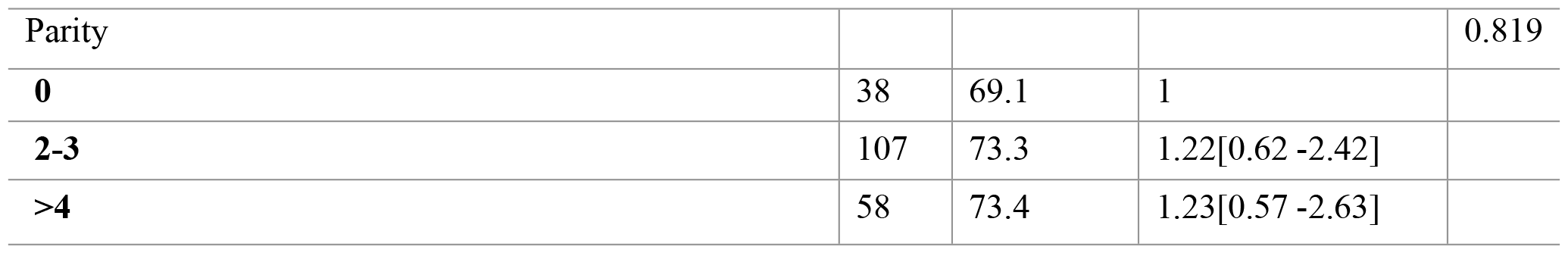
Association between socio-demographic characteristics and late start of ANC.

There was no statistically significant difference between women who started ANC early and those who started late in terms of age category, marital status, education of wife and husband, occupation of wife and husband, household size, gender and parity (P>0.05). No association was observed between these variables and the late start of ANC.

### Relationship between socio-cultural characteristics of pregnant womenand late start of ANC

**Table 5:** Association between socio-cultural characteristics and late onset of ANC.

| Variable | Late start of the ANC |  |  |  |
| --- | --- | --- | --- | --- |
|  | n | Percentage (%) | Crude OR (IC95%) | p. value |
| Media information (radio) |  |  |  | 0.236 |
| **No** | 87 | 76.3 | 1 |  |
| **Yes** | 116 | 69.9 | 0.72[0.41 -1.24] |  |
| Knowledge of ANC |  |  |  | 0.052 |
| **Better knowledge** | 87 | 66.9 | 1 |  |
| **Little knowledge** | 116 | 77.3 | 1.68[0.99 -2.86] |  |
| Benefits of doing ANC |  |  |  | 0.110 |
| **No** | 127 | 69.4 | 1 |  |
| **Yes** | 76 | 78.3 | 1.59[0.89 -2.84] |  |
| Decision by the woman about ANC |  |  |  | 0.321 |
| **Herself** | 152 | 71.0 | 1 |  |
| **Other (spouse, mother-in-law)** | 51 | 77.2 | 1.39[0.72 -2.64] |  |
| Encouragement from husband/ANC |  |  |  | 0.339 |
| **No** | 33 | 78.6 | 1 |  |
| **Yes** | **170** | **71.4** | **0.68[0.30 -1.50]** |  |

There was no statistically significant difference between women who started ANC early and those who started late on the variables of benefits of starting ANC early, women’s knowledge of the interventions offered during ANC, the person who made the decision to perform ANC and the husband’s support (encouragement) to perform ANC (p. value >0.05). There was no association between these factors and the delay in starting ANC.

### Relationship between pregnancy characteristics, those linked to the health care system and the delay in starting ANC

There was a statistically significant difference between women who underwent ANC early and those who underwent it late in terms of: the period during which the woman became aware of her pregnancy (p<0,001). Women who became aware of their pregnancy after 4 months initiated ANC later than others, pregnancy planning (p<0.009). Women who did not wish to be Pregnant start ANC later than others and time taken to get to the Health Facility (p<0.022) was also associated tp delay in starting ANC. Women living at a distance of more than 30 minutes’ walk from their home to the Health Facility underwent ANC later than women living at a distance of more than 30 minutes’ walk from their home to the Health Facility. Women who waited more than 30 minutes before being seen were late in undergoing ANC than those who did not.

. The table below shows the relationship between the characteristics of the pregnancy, those linked to the care system and the delay in starting ANC.

**Table 6:**
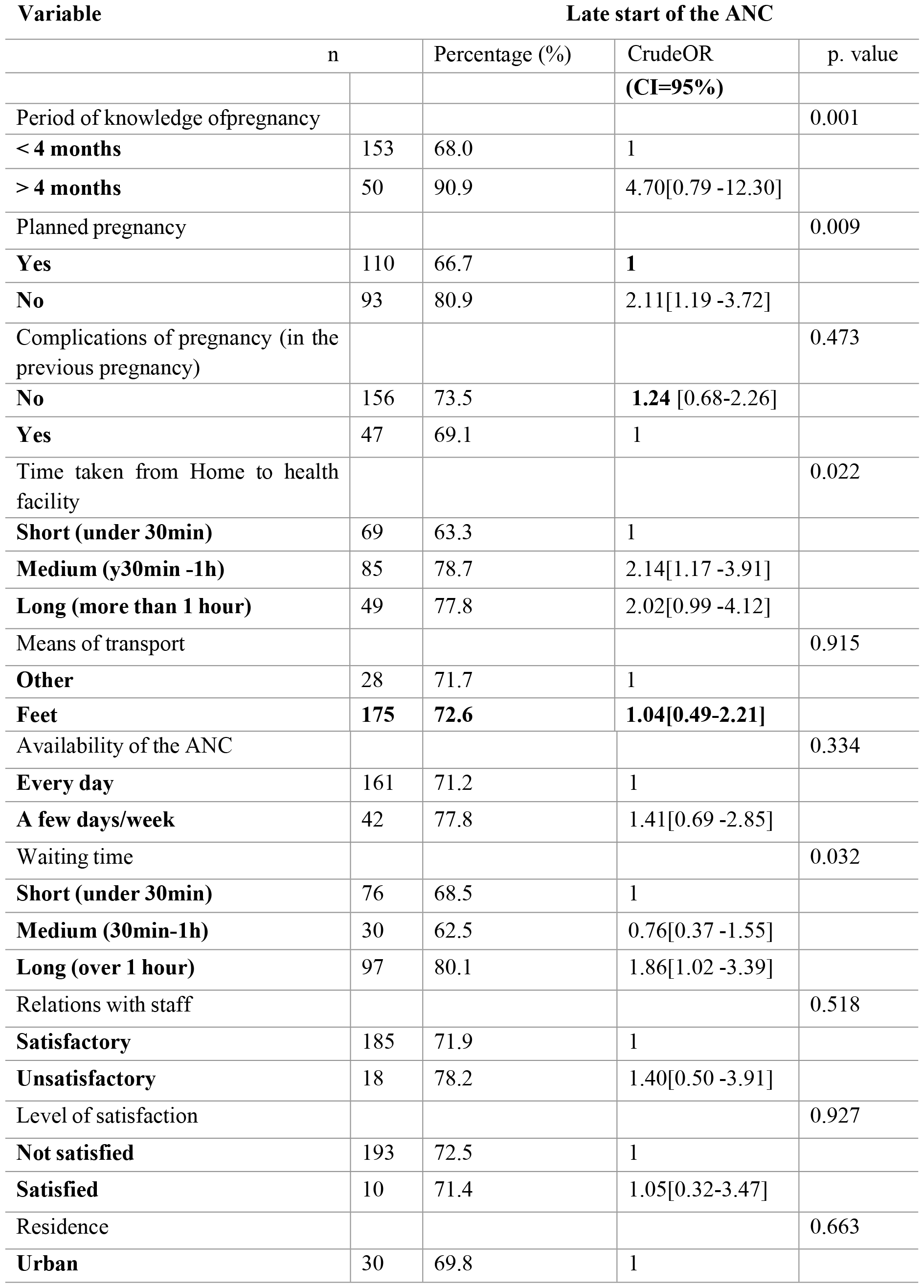
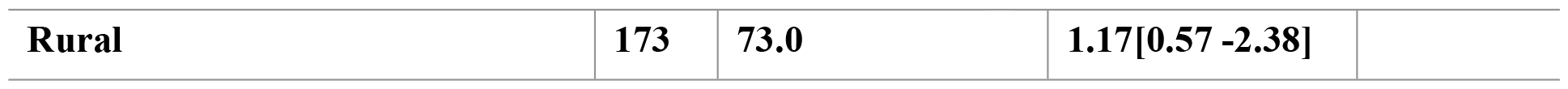
Association between pregnancy characteristics, characteristics of the healthcare system and delay in starting ANC.

### Factors associated with late start of ANC

The table 7 shows the variables statistically associated with late onset of ANC in a final multivariate logistic regression model. The variables that remained statistically significant in the final model were age, women’s knowledge of the interventions offered during ANC, the time at which the woman knew she was pregnant, the desire for pregnancy and the time taken to get to the Health Facility.

**Table 7:** Determinants of late start of ANC.

| Variables | Late start of ANC |  |  |  |
| --- | --- | --- | --- | --- |
|  | n | (%) | A OR [IC95%] | p-value |
| **Age** |  |  |  |  |
| Less than 20 years old | 19 | 90,5 | 1 |  |
| Between 20-35 years old | 149 | 70,3 | 0.18 [0.03–0.83] | 0.028 |
| Over 35 years old | 35 | 74,5 | 0.20 [0.03– 1.05] | 0.059 |
| **Knowledge of ANC** |  |  |  |  |
| Better knowledge | 116 | 66,9 | 1 |  |
| Little knowledge | 87 | 77,3 | 2.37 [1.32– 4.25] | 0.04 |
| **Period of knowledge of pregnancy** |  |  |  |  |
| < 4 months | 153 | 68,0 | 1 |  |
| > 4 months | 50 | 90,9 | 6.70[2.41– 18.66] | < 0.001 |
| **Planned pregnancy** |  |  |  |  |
| Yes | 110 | 66,7 | 1 |  |
| No | 93 | 80,9 | 1.85 [1.01– 3.40] | 0.047 |
| **Time taken from Home health facility** |  |  |  |  |
| Short (under 30min) | 69 | 63,3 | 1 |  |
| Medium (30min -1h) | 85 | 78,7 | 2.40 [1.25– 4.59] | 0.008 |
| Long (more than 1 hour) | 49 | 77,8 | 2.15 [1.01– 4.60] | 0.047 |

Women aged 20-35 years had an 82% lower risk of starting ANC late than teenagers (AOR = 0.18; 95%CI:0.03-0.83). Women aged over 35 years old had a lower risk of starting ANC late than women under 20 years. Women with poor knowledge of the interventions offered by the ANC service were 2.37 timesmore likely to start ANC late than women with better knowledge (AOR = 2.37; 95%CI:1.32-4.25).

Pregnant women who knew they were pregnant after 4 months were around 7 times morelikely to start ANC late than those who knew they were pregnant before 4 months (AOR= 6.7; 95%CI:2.41-18.66), and those who had unplanned pregnancies were 1.85 times more likely to start ANC late than those who had planned their pregnancies (AOR = 1.85 95%CI:1.01-3.40)

The time taken by women to get to the health facility from home is associated with the delay in starting ANC. Women who walk between 30 minutes and 1 hour and more than an hour have a risk of starting ANC late that is 2.40 and 2.15 times higher respectively than women who get to the health facility in less than 30 minutes ((AOR = 2.40 95%CI:1.25-4.59), (AOR = 2.15; 95%CI:1.01-4.60).

## DISCUSSION

Early initiation of ANC and regular visits have a positive influence on maternal and fetal outcomes(12). However, the results of this study show that women undergo ANC late and do not fully benefit from all the interventions offered during ANC. The delay in performing ANC has been noted by other authors around the world. The frequency differs between developed and developing countries. It is low in wealthy countries, but remains higher in low- and middle-income countries(8). Thus the frequency of women initiating ANC late after 12 weeks of pregnancy represents less than 20% in developed countries (13) (14), (15) (16).

On the other hand, this frequency exceeds 50% in developing countries. This observation was made in Ethiopia and Mynamar (17)(18).In the East African region to which Burundi belongs, the onset of ANC is also late, beyond 12 weeks, as noted in our study, and the frequency exceeds 70% according to the study carriedout in Kenya, Rwanda, and Tanzania (19), (20) (21).Delays in initiating antenatal care have an impact on the coverage of ANC required during pregnancy. A study carried out in France in 2017 showed that performing less than 50% of the recommended ANC is associated with severe maternal morbidity and severe perinatal morbidity(15). Gestational age at the start of the first antenatal visit ranged from 1 month (4 weeks) to 9 months (40 weeks) of pregnancy. The majority of women who undergo late ANC do so in the second trimester. Our results are similar to those found in Zambia (22), Ethiopia (23), Nigeria (24), Ghana (25)and South Africa(26). The lack of information or sufficient knowledge about the importance of starting ANC in the first trimester would explain this delay.

A study exploring the factors influencing use of antenatal care in four sites in three sub-Saharan countries (Kenya, Malawi and Ghana) found that in Kenya and Malawi, women were warried of attending antenatal care because they would be informed of their HIV status. Husbands often refused to be tested and, in the most extreme cases, instead accused their wives of adultery and abandoned them. This had implications for women’s participation and delay in antenatal care(27). Another study carried out in Tanzania suggests that fear of HIV disclosure, socio-cultural beliefs about pregnancy such as witchcraft, lack of support from the partner and lack of respect from providers towards the pregnant woman were the main barriers to early antenatal care(28). The lack of knowledge about the importance of early visits, previous births without complications, fear of shame and stigmatization of pregnancies outside marriage or among adolescents, and cultural beliefs about pregnancy are the main obstacles to early ANC consultation(29). In Rwanda, lack of knowledge about the benefits of early ANC, poverty and problems related to lack of health insurance(30), and the requirement for the spouse to be present during ANC (31)are the barriers to early initiation of ANC mentioned by women.

Pregnancy in young women under the age of 20 (adolescents) is a factor associated with delayed onset of ANC. Women aged 20-35 had a lower risk of starting ANC late than younger women aged under 20.Similar results have been found in Zambia (26).Teenage girls lack information about the right time to start ANC. They often have unplanned pregnancies because some do not know how to avoid pregnancy, while others are unable to obtain contraception, refuse unwanted sex or resist forced sex. Women with poor knowledge of the interventions offered by the ANC service were more likely to start ANC late than women with better knowledge. This finding has been noted by other authors, in studies carried out in Ethiopia (23), (33) and Zambia (34).Women who do not know about the interventions offered during ANC do not detect or are not even aware of the importance of ANC for the health of the mother and the fetus and are unaware of the correct time to start ANC, It has been shown that women who received information about antenatal care, pregnancy risks and danger signs were more likely to start ANC early than women who received no information about these situations(35). This implies that women’s knowledge of the services offered during ANC, the danger signs and the advantages of performing ANC early would stimulate pregnant women to initiate ANC early and probably also to complete the required number of ANC.

Women who were aware of their pregnancy after 4 months were more likely to start ANC late than those who were aware of their pregnancy before 4 months Similar results have been found in Ethiopia (36). the study conducted in southern Tanzania, indicate late knowledge of one’s pregnancy status as a good predictor of delayed initiation and participation in antenatal care(37). This delay in becoming aware of one’s pregnancy status could be explained by women who have used hormonal contraceptives, particularly injectables, and found themselves pregnant without having menstruated after stopping the contraception and, women who have had close pregnancies. Women who had not planned their pregnancy delayed starting ANC more than those with a planned pregnancy. Non-planning of pregnancy is a predictor of late onset of ANC, which has been consistently found in South Africa (26) and Ethiopia (33) (38,39), Zambia (34)and Rwanda(20).

Women who have not planned their pregnancy lack support from their partner or family, and this contributes to the delay in initiating antenatal care. Another reason would be that women who have had pregnancies at an unplanned time refuse to be in this state and try to hide their pregnancy, which hurts the delay in starting ANC. A study conducted in South Africa reported that some women who had not planned their pregnancy considered terminating it (26)and it was adolescents who were more likely to have unplanned pregnancies and thus late for their first ANC. The time taken by women to get from their home to the health facility is associated with the delay in starting ANC. Similar results was found in Ethiopia (38,40).

The time taken from home to the health facility is an important predictor of the delay in starting ANC. The results of the study showed that 84.64% of the women surveyed lived in rural areas. As the Muramvya district is located in the geographically mountainous Mugamba natural region, travel is mainly on foot or by motorbike, and the latter are not financially accessible to everyone. As a result of these travel difficulties, women take longer to walk before arriving at the health facility and consequently delay starting their first ANC.

### Limitations and strengths of the study

As the study only included women attending health facilities, it was not possible to determine the prevalence of late-start ANC in the district, it was not possible to obtain information from women who did not undergo ANC. There could have been a memory bias for gestational age, although verified in the mother-child record, had been determined based on the woman’s recall of the date of her last menstrual period. Given that the study took place in health facilities, it is possible that factors concerning the relationship between the woman and the health care staff and the woman’s level of satisfaction with the services offered during ANC were under-reported.

## CONCLUSION

The results of the study reveal that the proportion of women who initiate ANC late remains high despite the availability of antenatal care in all the public and faith-based health facility in the district. Young maternal age and lack of sufficient knowledge about ANC services are observed socio-demographic and cultural factors associated with late onset of ANC. Among the factors related to pregnancy and accessibility to antenatal care services, late knowledge of pregnancy, unplanned pregnancy and the time taken to get to the Health Facility were found to be associatedwith late start of ANC. Strategic actions aimed at paying particular attention to teenage pregnancies and improving women’s level of knowledge about the interventions offered by the ANC service, and access to information about family planning methods, could reduce the number of women starting ANC late.

## Data Availability

Data are available

## Acknowledgments

The authors would like to express their gratitude to all women who participated in the study by responding and sharing their opinions to the research team. We would also like to thank our fieldworkers who participated actively in data collection.

## Author contributions

Conceptualization : Jean Marie Butoyi

Data curation : Jean Marie Butoyi

Formal analysis : Jean Marie Butoyi

Funding acquisition : Jean Marie Butoyi

Investigation : Jean Marie Butoyi

Methodology : Jean Marie Butoyi,

Software : Jean Marie Butoyi

Supervision : Anatole Nkeshimana, Yves Coppieters

Writing-original draft: Jean Marie Butoyi, Caleb Kimutai Sagam, Belyse Munezero

Writing-review &editing: Jean Marie Butoyi, Caleb Kimutai Sagam, Belyse Munezero

